# Whole Genome Sequencing to Study SARS-CoV-2 Transmission between University Students and the Surrounding Community in Pittsburgh, Pennsylvania, 2020-2021

**DOI:** 10.1101/2025.10.14.25337767

**Authors:** Vatsala Rangachar Srinivasa, Marissa P. Griffith, Kathleen A. Shutt, Hunter Coyle, Nathan J. Raabe, Kady D. Waggle, Tung Phan, Graham M. Snyder, Lora Lee Pless, Daria Van Tyne, Alexander J. Sundermann, Elise M. Martin, Lee H. Harrison

## Abstract

SARS-CoV-2 transmission was investigated between university students and the surrounding community using whole genome sequencing. Fourteen putative transmission clusters were identified. Proximity assessed using ZIP codes showed that clustered cases were more widely dispersed than non-clustered cases, highlighting the need for integrated surveillance, coordinated interventions, and data-driven public health policies.

## Introduction

Between June 2020–April 2021, the highest nationwide incidence of COVID-19 in the U.S. was among 18–29-year-olds, which represents most university students [1]. Even though students are less likely to develop severe COVID-19, identifying transmission is vital for protecting susceptible faculty, staff, and the surrounding community [1-3]. Using whole genome sequencing (WGS), here we investigated SARS-CoV-2 transmission between University of Pittsburgh students and the surrounding community.

## Methods

### Study Setting

SARS-CoV-2 specimens were collected between July 2020–April 2021 from students attending the University of Pittsburgh’s sprawling urban main campus (detailed in [4]), and from patients and healthcare workers (HCWs) at 12 regional hospitals within an integrated healthcare system across the greater Pittsburgh region. Ethics approval was obtained from the University of Pittsburgh’s Institutional Review Board (STUDY22120012).

We assumed that SARS-CoV-2 specimens from the hospital population were representative of the community surrounding the university campus. Rationale for sequencing these specimens has been detailed previously [5, 6]. Specimen collection from students, laboratory methods, and bioinformatics analyses were previously described [1].

### Genomic data analysis

We used a pairwise SNP cutoff of 2 with average linkage hierarchical clustering to identify genetically related clusters [1, 5]. FastTree (v0.11.2, default parameters) was used to construct a maximum likelihood phylogenetic tree based on the generalized time-reversible (GTR) model.

### Epidemiological analyses

To evaluate potential transmission directionality and overlap between students and community members, we defined a putative transmission cluster as containing genetically related SARS-CoV-2 genomes belonging to both population groups. We then assessed the relationship between geographic proximity and clustering using ZIP code data. Specimens were excluded if the ZIP code was missing. Patient names and demographic information were reviewed to remove duplicate specimens across two groups.

To ensure appropriate geographic assignment, community SARS-CoV-2-positive specimens were stratified into two groups: i) Ambulatory Community: included outpatients (tested in an outpatient clinic, emergency department, or within three days of hospital admission) and HCWs. For this group, we used residential ZIP codes as geographic reference; ii) Hospitalized Community: included inpatients who tested positive ≥3 days after hospital admission, suggesting healthcare-associated infection. Therefore, the hospital where the specimens were collected served as the geographic reference. For students, addresses reported during the university’s pandemic contact tracing efforts were used to obtain ZIP codes for where they were residing.

Pairwise distances between ZIP codes of students and community members were calculated using the Haversine formula (geosphere package in R). Mann-Whitney U test was used to assess whether geographic proximity was associated with clustering among genomes belonging to the same SARS-CoV-2 lineage across the two population groups.

## Results

We identified 664 SARS-CoV-2 specimens from 653 individuals after excluding 16 with missing ZIP codes and two that failed WGS quality control (≥95% coverage at 10× depth). Of these, 264 (39.8%) were from students and 400 (60.2%) were from community members (**Supp. Table 1**); 339 (51%) specimens were collected during the fall semester of 2020 and 325 (49%) in the spring semester of 2021 (**Supp. Figure 1A and 1B**). Among community specimens, 299 (74.8%) belonged to the Ambulatory Community (269 outpatients and 30 HCWs), while 101 (25.3%) belonged to the Hospitalized Community (**Table 1**). Community members were predominantly female (231, 59.4%) and white (287, 73.8%), with ages ranging from 9 months to 98 years (median=54 years).

**Table 1.** Demographic characteristics of community members.

| **Characteristic**<br># Patients (%) | **Overall**<br>N = 389 | **Ambulatory Community** |  | **Hospitalized Community**<br>N = 91<br>(23.4%) | **p-value** |
| --- | --- | --- | --- | --- | --- |
|  |  | **Outpatient**<br>N = 268<br>(68.9%) | **Employee**<br>N = 30<br>(7.7%) |  |  |
| **N specimens** | 400 | 269 | 30 | 101 |  |
| **Sex** |  |  |  |  | 0.003 |
| Female | 231 (59.4%) | 158 (59.0%) | 26 (86.7%) | 47 (51.7%) |  |
| Male | 158 (40.6%) | 110 (41.0%) | 4 (13.3%) | 44 (48.4%) |  |
| **Race** |  |  |  |  | 0.12 |
| White | 287 (73.8%) | 192 (71.6%) | 25 (83.3%) | 70 (76.9%) |  |
| Black | 77 (19.8%) | 60 (22.4%) | 4 (13.3%) | 13 (14.3%) |  |
| Unknown | 19 (4.9%) | 10 (3.7%) | 1 (3.3%) | 8 (8.8%) |  |
| Other | 6 (1.5%) | 6 (2.2%) | 0 (0%) | 0 (0%) |  |
| **Age** |  |  |  |  | <0.001 |
| Median [SD] | 54 [20.5] | 52 [19.7] | 34 [12.1] | 67 [17.0] |  |

**Figure 1A.**
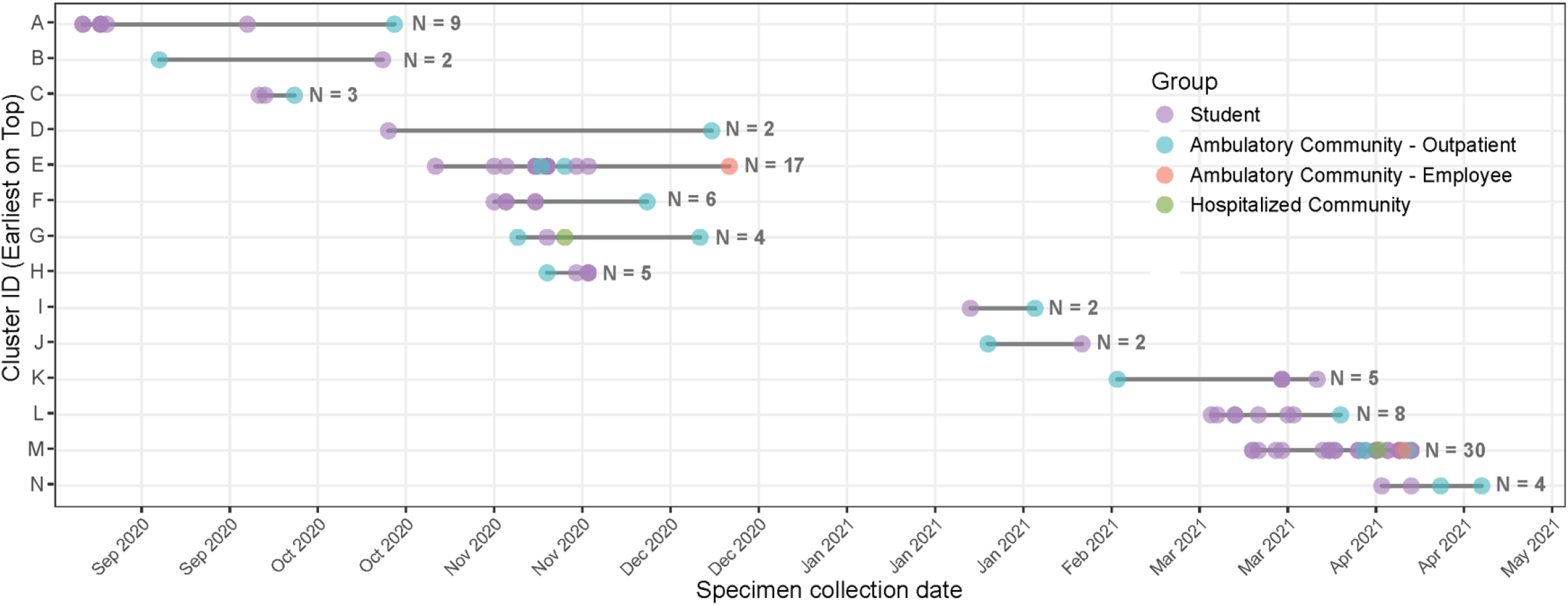
Putative transmission clusters among students and the surrounding community. Each data point represents a sequenced SARS-CoV-2 specimen in a cluster, colored by demographic groups. The horizontal line represents the cluster duration. N indicates the number of patients within each cluster. Visualization created in R using ggplot2 package. Important dates: Fall term move-in days, 07/15/2020 – 08/19/2020; Family weekend, 09/25/2020; Winter recess and spring term move-in days, 12/06/2020 – 01/19/2021; St. Patrick’s Day, 03/17/2021.

We identified 52 SARS-CoV-2 lineages, with B.1.2 (254, 38.3%) and B.1.1.7 (134, 20.2%) predominating. Seven lineages containing 18 genomes were unique to students, 16 (88.9%) of which coincided with semester move-in days, St. Patrick’s Day, winter recess, or family weekend (09/25/2020─09/27/2020), and were supported by contact tracing links [1]. There were 23 lineages with 54 genomes unique to community patients, ranging from 1─14 patients/lineage. Same-lineage, pairwise SNP distances ranged from 0─52, with most genetic relatedness explained within 2-SNPs, thus supporting our choice of a 2-SNP threshold for clustering (**Supp. Figure 1C**).

**Figure 1B.**
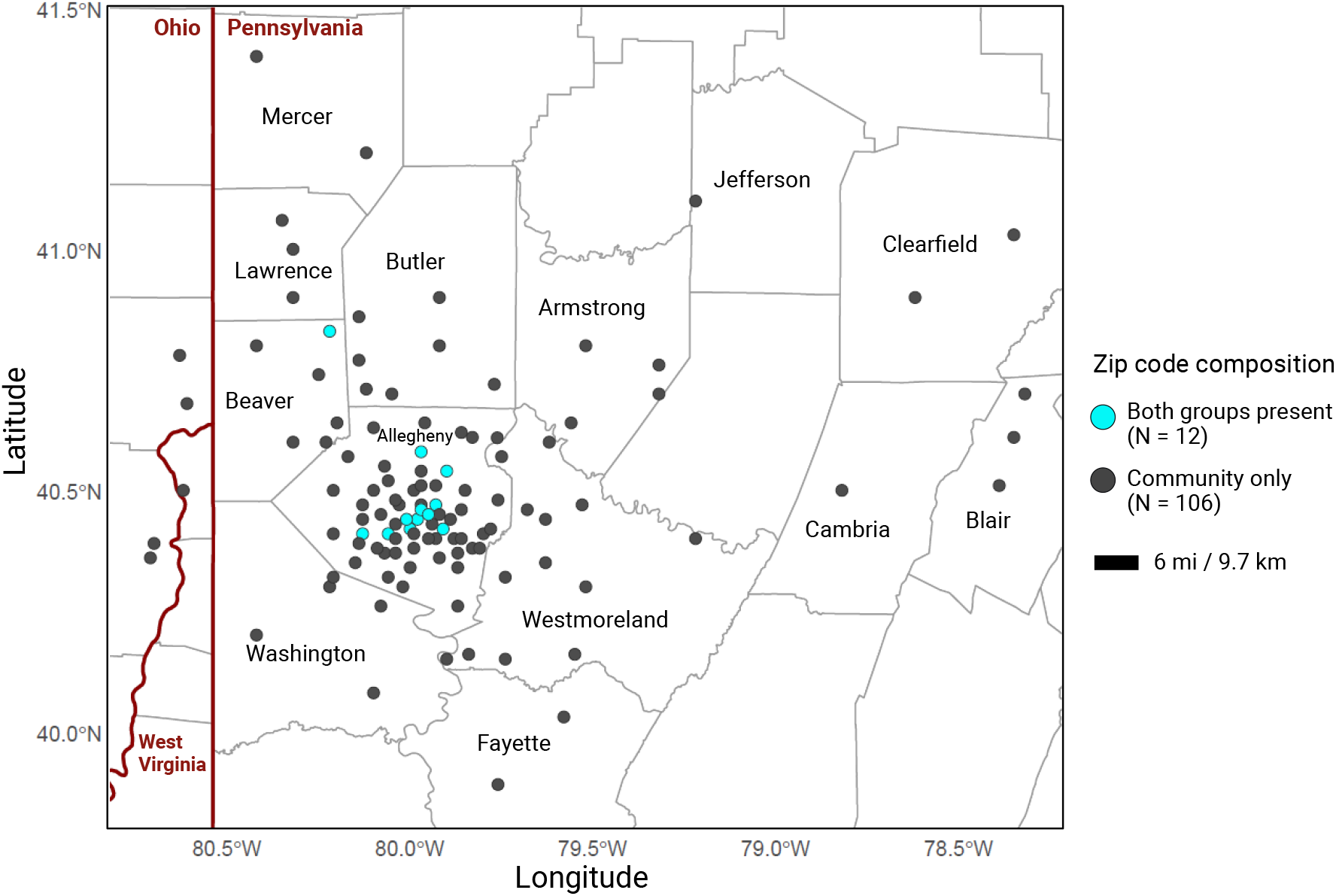
Spatial distribution of ZIP codes from which SARS-CoV-2 specimens were collected. The geographic borders represent the state (dark red) and county (grey) boundaries. Each data point represents a ZIP code that contributed SARS-CoV-2 specimens. N indicates the number of ZIP codes represented by each group.

**Figure 1C.**
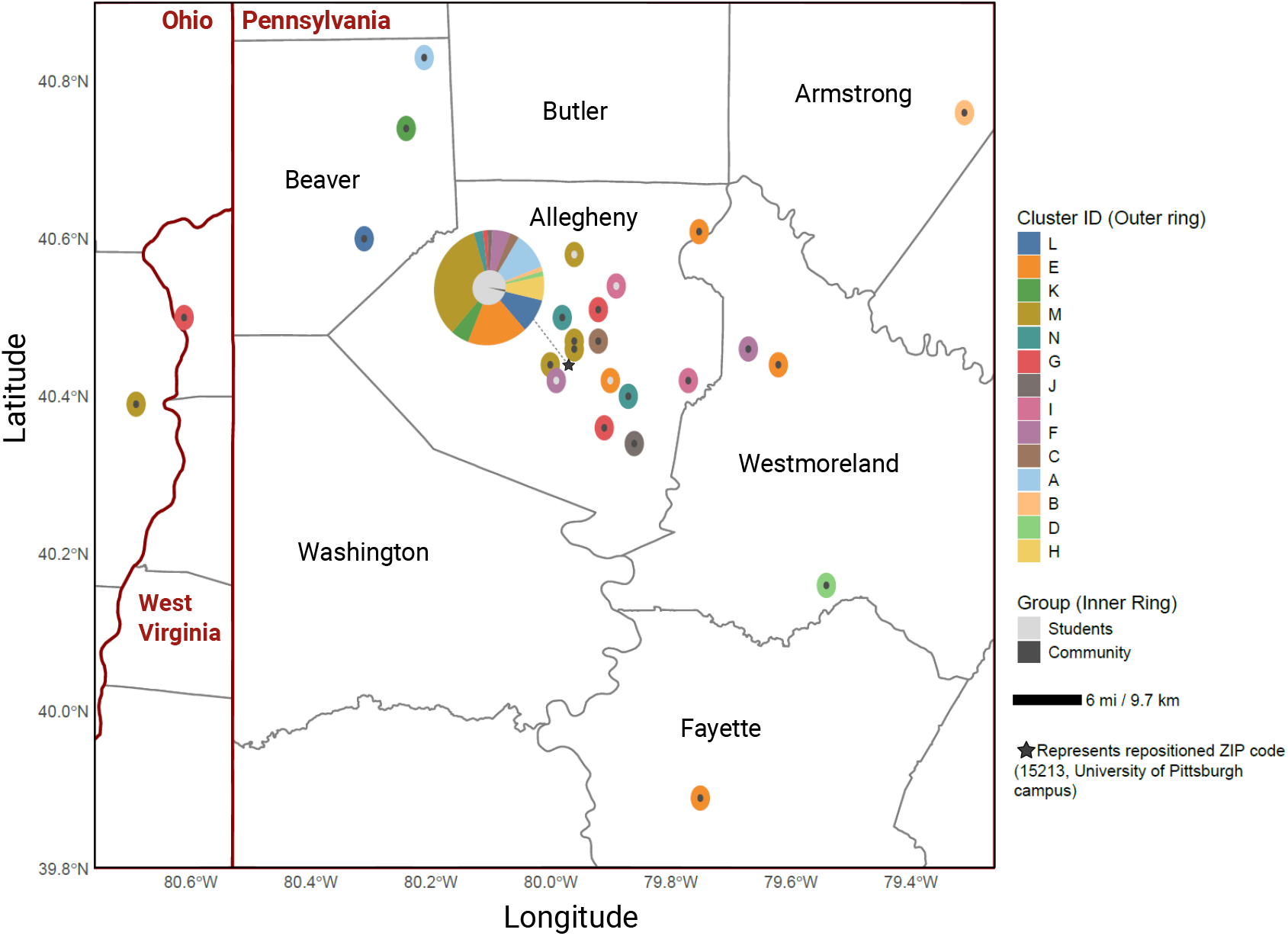
Spatial distribution of putative transmission clusters. The outer ring represents the clusters, and the inner ring represents specimen grouping. Pie charts indicate ZIP codes containing multiple clusters and/or specimen groups. The geographic borders represent the state (dark red) and county (grey) boundaries. Visualization created in R using tigris and ggplot2 packages.

Overall, 99/664 (15%) genomes were closely genetically related, forming 14 putative transmission clusters (range=2─30 genomes/cluster), each involving students and community members. Of these, 77 (77.8%) were from students, and 22 (22.2%) from community members (**Supp. Table 2**). The average cluster duration was 28 days (range=6─55 days). In nine clusters (64.3%), the earliest specimen was from a student, while the remaining five began in the community (**Figure 1A**). Fewer clusters were noted during Dec 2020─Jan 2021 when students were on winter break.

Specimens were collected from 110 ZIP codes across 17 counties, reflecting a broad geographic distribution (**Figure 1B**). Putative transmission clusters were widely dispersed, with a median pairwise geographic distance of 15 miles (mean=15.5; range=0─41; **Figure 1C**). In contrast, non-clustered cases had a median distance of 5.5 miles (mean=10.8; range=0─171; p=0.0007).

## Discussion

WGS surveillance provided valuable insights into SARS-CoV-2 transmission dynamics between students on an urban campus and the surrounding community. We identified 14 overlapping clusters between the two groups, most (64.3%) originating from students. Interestingly, the median distance for clustered cases was 15 miles vs. 5.5 miles for non-clustered cases, indicating that putative transmission extended beyond the immediate campus area and contributed to a broader regional spread. These findings underscore the difficulty in preventing transmission despite multiple interventions implemented by the university [4].

We observed that 13% of lineages occurred only among students, whereas 44% were unique to the community, indicating greater lineage diversity among community members, consistent with prior findings [7]. Furthermore, 42% of lineages were shared between students and the community, indicating substantial overlap in circulating lineages. However, this may also suggest that some circulating lineages were not sequenced, highlighting inherent limitations of convenience sampling.

Prior studies in the U.S. using publicly available genomes have shown clustering between SARS-CoV-2 sampled from students and the local community [3, 8, 9]. Availability of ZIP code data strengthened our study by enabling assessment of the relationship between geographical proximity and clustering, which is relevant in a city like Pittsburgh, PA that has over 20 colleges and universities interspersed throughout the community. Prior studies have shown that geographically intertwined population groups foster cross-transmission [10], and medical students may act as bridges for transmission to the community [7]. It is also possible that students frequently visit nearby family and friends, contributing to the spread, potentially explaining why clustered specimens had a narrower distance range (0─41 vs. 0─171 miles) but a higher median distance than non-clustered cases. However, we did not identify student degree programs and did not have data beyond ZIP codes to establish epidemiological links.

Our study had several limitations. We sequenced a small subset of the community and student specimens, which may not reflect cases that were not collected and/or not sequenced. Community specimens were sampled from hospitals, which could underrepresent viral transmission and diversity. ZIP codes were used as a proxy for geographic proximity, which offers relatively low geographic resolution. We inferred transmission based on ZIP codes, but transmission could have occurred elsewhere. Some clusters may represent widely circulating community strains that meet the stringent 2-SNP cut-off without indicating true epidemiological links. Finally, the study was conducted in a single university─community setting and, therefore, the findings may not be generalized.

In conclusion, this hypothesis-generating study suggests that SARS-CoV-2 transmission between University of Pittsburgh students and the surrounding community was substantial, yet complex, underscoring the need for integrated genomic surveillance, coordinated intervention strategies, and data-driven public health policies.

## Data Availability

The NCBI, GISAID and SRA accession numbers for the respiratory viral genomes used in this study can be found in Supp. Table 1. Sequence data can be found at NCBI BioProject PRJNA1337675.

## Acknowledgments

We are grateful to Ms. Melissa McCullough, UPMC Clinical Microbiology/Virology lab, for helping with SARS-CoV-2 specimen collection. We also appreciate Dr. Jane Marsh’s support in conceptualizing the study design.

## Data Sharing Statement

The NCBI, GISAID and SRA accession numbers for the respiratory viral genomes used in this study can be found in **Supp. Table 1**. Sequence data can be found at NCBI BioProject PRJNA1337675.

## Author Contributions

VRS: Conceptualization, Methodology, Analysis, Data Curation, Writing - Original Draft, Writing - Review & Editing, Visualization

MPG: Data Curation, Analysis, Writing - Review & Editing

KAS: Data Curation, Writing - Review & Editing

HC: Data Curation & Visualization, Writing - Review & Editing

NJR: Data Curation & Visualization, Writing - Review & Editing

KDW: Data Curation, Writing - Review & Editing

JWM: Conceptualization, Writing - Review & Editing

GMS: Conceptualization, Writing - Review & Editing

LLP: Conceptualization, Resources, Project administration, Writing - Review & Editing

DVT: Conceptualization, Methodology, Supervision & Writing - Review & Editing

AJS: Conceptualization, Methodology, Analysis, Supervision, Writing - Review & Editing

EMM: Conceptualization, Methodology, Data Curation, Supervision, Funding acquisition, Writing - Review & Editing

LHH: Conceptualization, Methodology, Analysis, Resources, Data Curation, Writing – Original

Draft, Writing - Review & Editing, Supervision, Project administration & Funding acquisition

---

**Supplementary Figure 1A.**
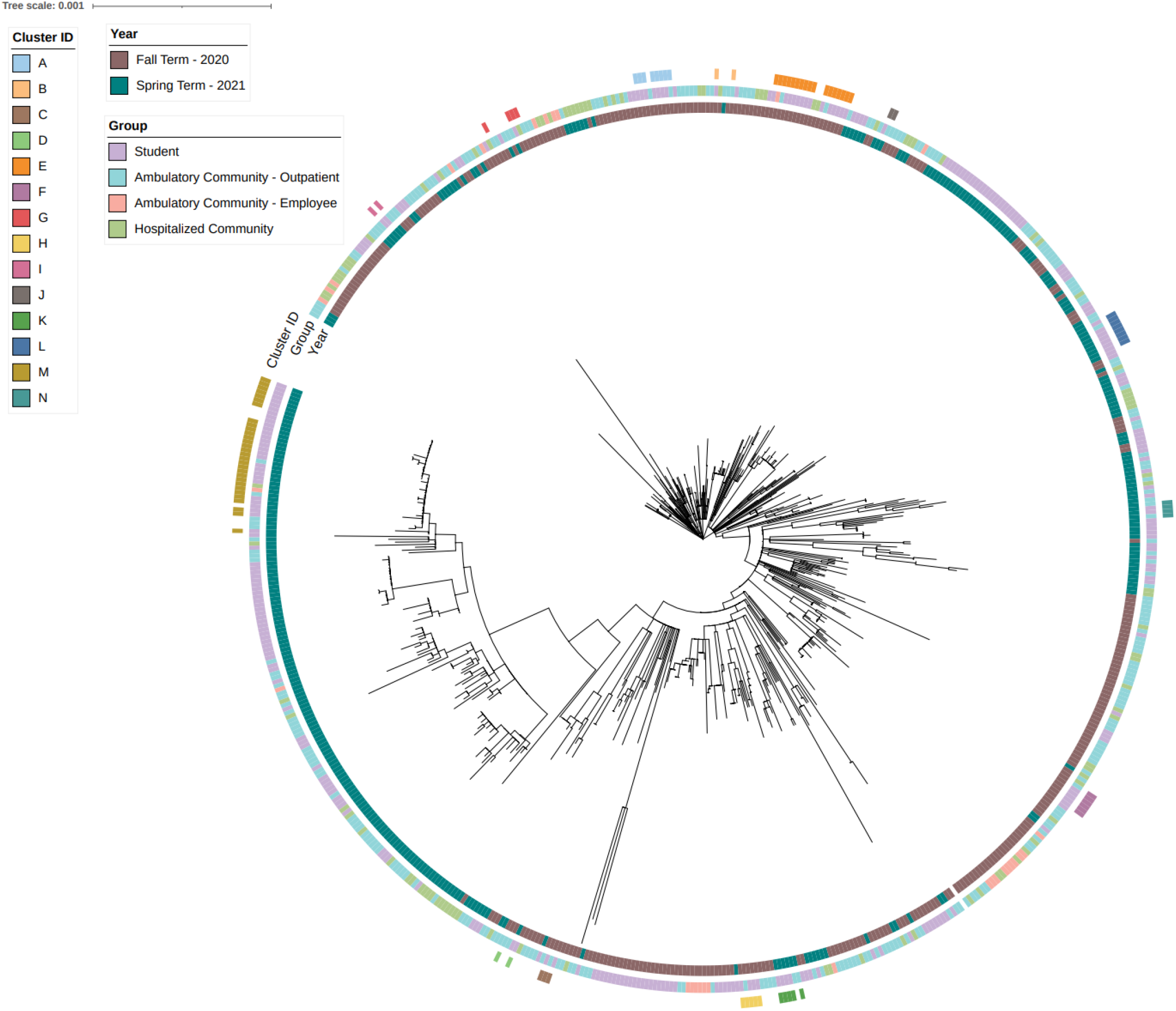
Phylogenetic tree of SARS-CoV-2 genomes. Tree scale represents the nucleotide substitutions per site. Vertical strips represent specimen collection year, specimen grouping, and putative transmission clusters. Visualization created on iTOL.

**Supplementary Figure 1B.**
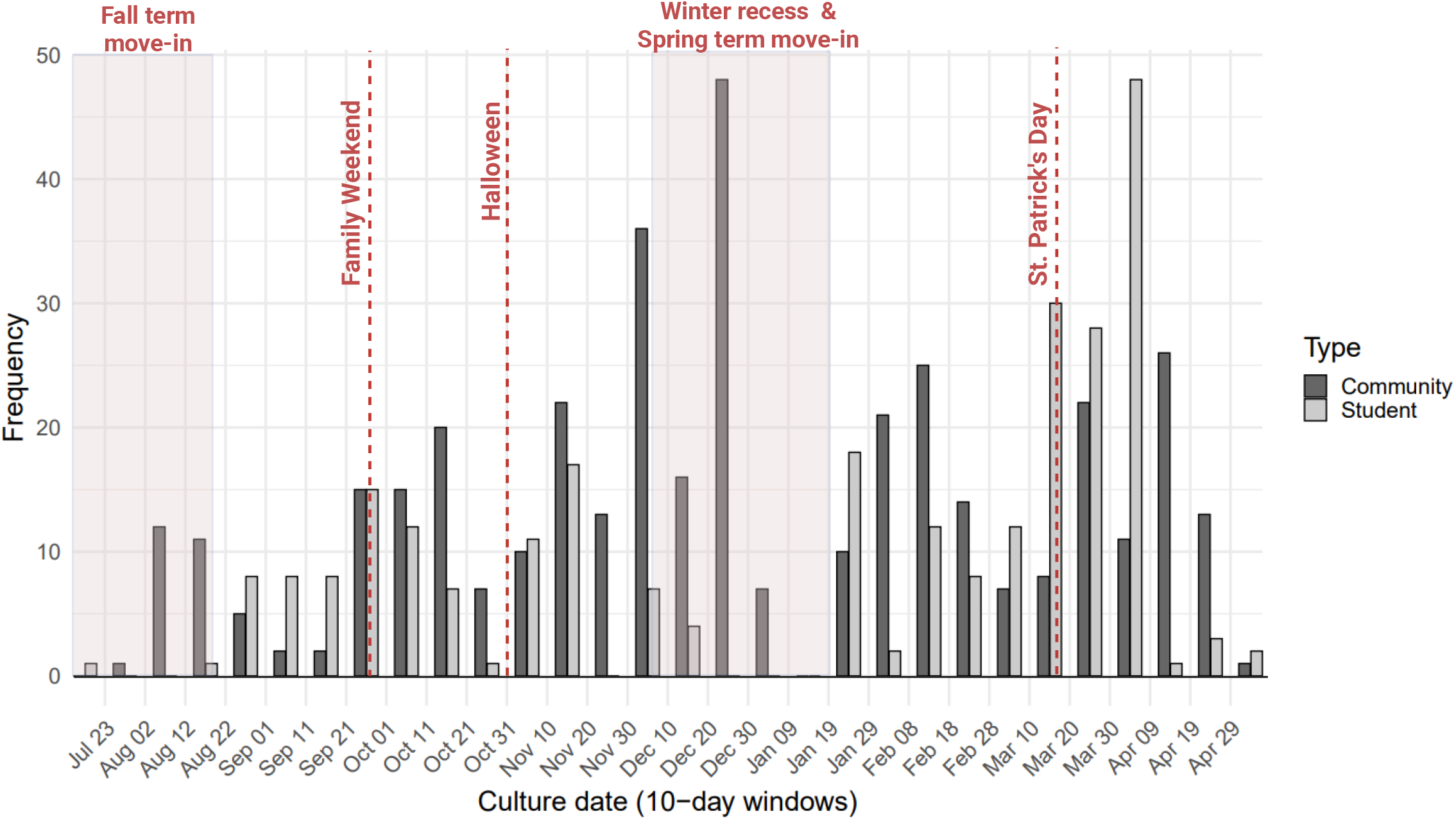
Distribution of SARS-CoV-2-positive specimens that were sequenced. Dark red dashed lines and boxes indicates: Fall term move-in days, 07/15/2020 – 08/19/2020; Family weekend, 09/25/2020; Winter recess and spring term move-in days, 12/06/2020 – 01/19/2021; St. Patrick’s Day, 03/17/2021.

**Supplementary Figure 1C.**
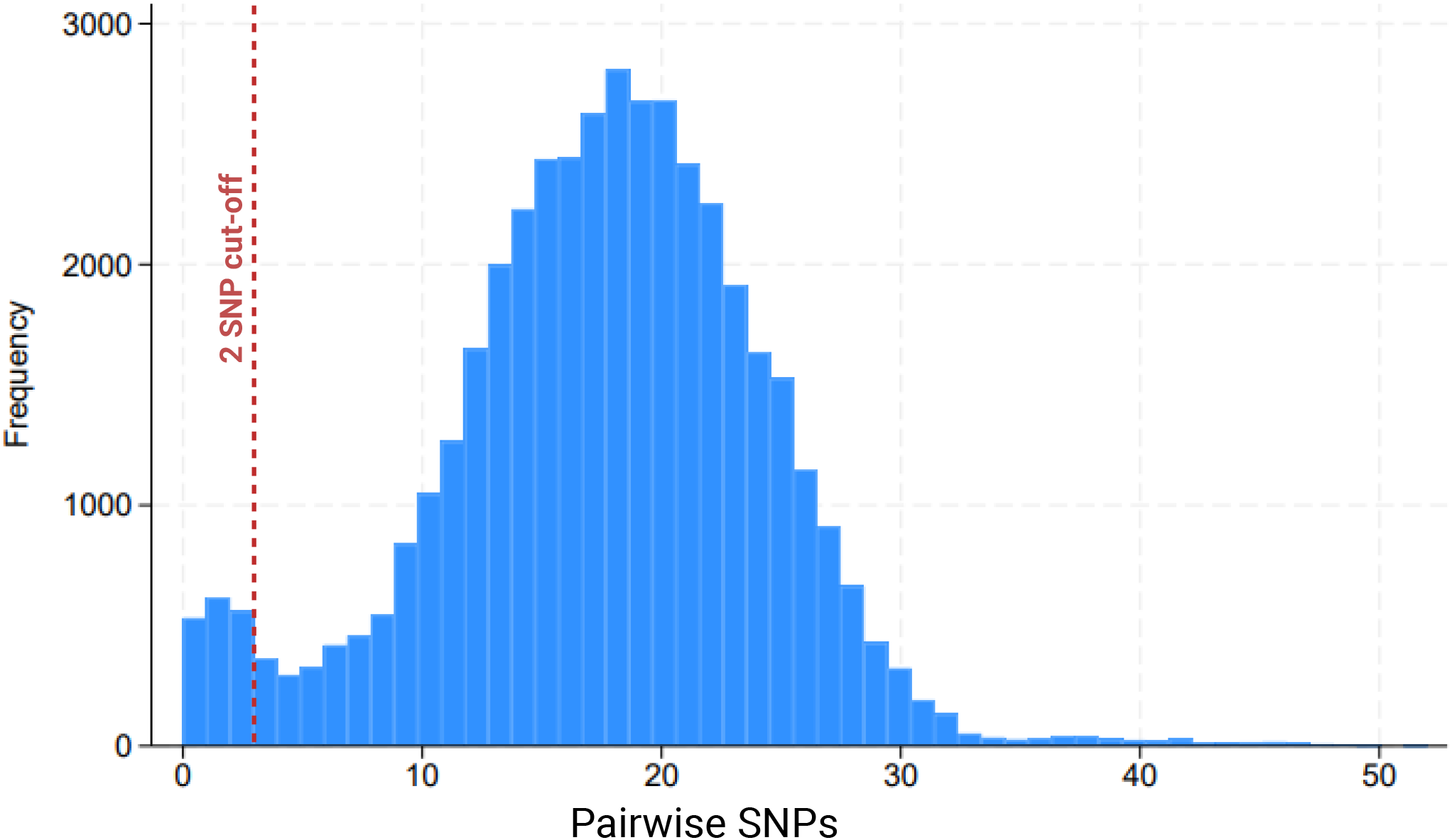
Distribution of Pairwise SNP distances of SARS-CoV-2 genomes belonging to the same lineage. Dark red dashed line indicates 2 SNP threshold.

